# Optimized multichannel tDCS protocol for clinical use in patients with major depressive disorder

**DOI:** 10.1101/2025.02.12.25322157

**Authors:** Mohammad Ali Salehinejad, Marzieh Abdi, Mohsen Dadashi, Amir-Homayun Hallajian, Kiomars Sharifi, Ali Khadem, Ricardo Salvador, Giulio Ruffini, Michael A. Nitsche

## Abstract

Application of transcranial direct current stimulation (tDCS) has been increased in neuropsychiatric disorders, especially depression, in the last decade. Despite promising results, the clinical efficacy of tDCS is still under debate and researchers and clinicians try to maximize efficacy by developing optimized/individualized protocols. In this randomized sham-controlled, parallel-group clinical trial, we optimized a stimulation protocol for increasing and decreasing activity in the left and right dorsolateral prefrontal cortex (DLPFC). Sixty patients with unipolar depression received 30 sessions of conventional, optimized multichannel, or sham tDCS in parallel groups. In the multichannel optimized tDCS, electrical current was delivered via 7 small electrodes while in the conventional and sham tDCS large anodal and cathodal electrodes were used. Intervention efficacy and treatment response were evaluated before treatment and after the 10^th^, 20^th^, 30^th^ sessions, and 1- and 3-months post-intervention. Cognitive functions and brain connectivity were also assessed. Compared to the sham group, both active tDCS groups significantly reduced depressive symptoms up to 3 months following the intervention, and the therapeutic effects were observed from session 10^th^ only in the multichannel tDCS groups. Furthermore, the effect size from the optimized multichannel protocol was significantly larger than the conventional tDCS. These data suggest that optimized multichannel tDCS might maximize therapeutic efficacy in major depression.

## 1 Introduction

Major depression (MD) is the most common mental health disorder, affecting about 350 million people worldwide ^1^. It is the second most frequent disease with the highest age-adjusted disability, according to the latest report about the global burden of disability ^2^. Individuals with MD are affected by depressed mood, diminished interests, impaired cognitive function, and disturbed sleep or appetite ^3^. Antidepressants, especially Selective Serotonin Reuptake Inhibitors (SSRIs), and psychotherapeutic interventions, especially cognitive-behavioral therapy, are considered first-line treatments for MD with severe and mild to moderate severity, respectively ^4,5^. These treatments, nevertheless, come with some limitations. For example, SSRIs have delayed therapeutic benefits, taking about 4 to 6 weeks to ameliorate symptoms ^6^. Psychotherapy is mostly beneficial for patients with mild and moderate illness severity, but more than half of MD patients receiving therapy do not respond, and only one-third achieve remission. The latest meta-analysis of psychotherapy outcomes for eight mental disorders indicates that while psychotherapies are effective compared to control conditions, the absolute response rates are modest, suggesting a need for more effective interventions for patients who do not respond to first-line treatments ^7^. Accordingly, researchers and clinicians have been interested in developing other potential therapeutic interventions that do not have such limitations.

No established mechanism can explain all aspects of MD. However, previous neuroimaging studies have shown that MD is associated with functional changes in brain circuits such as the cognitive control network and the affective salience network ^3,8,9^. Furthermore, studies that monitored resting state electroencephalography (EEG) of patients with depression have shown that depression is associated with generally reduced and/or altered neural synchronization patterns ^10,11^ and lower coherence ^11,12^ in the default mode and central executive networks which are related to both cognitive impairment and emotional dysregulation in depression. These alterations in neural networks are linked to negative thinking patterns and cognitive symptoms of depression including abnormal self-referential thinking and mind-wandering (shown as rumination in depression) ^13^. Importantly, they provide opportunities for developing novel interventions that aim to more specifically restore impaired functional changes in MD.

Noninvasive brain stimulation techniques provide unique opportunities to not only study brain functions but also to modify core physiological parameters of human behavior and cognition (e.g., neuroplasticity) in both healthy and clinical populations ^14,15^. The success of some noninvasive brain stimulation techniques, such as repetitive transcranial magnetic stimulation (rTMS) – approved by the Food and Drug Administration (FDA) for the treatment of MD ^16^ – suggests that other techniques with related physiological effects may be considered as a potential intervention for patients with MD. Transcranial direct current stimulation (tDCS) is a non-invasive brain stimulation technique that uses a weak direct electrical current to modulate brain activity and excitability ^17,18^ with a safe profile ^19,20^. The exact mechanisms by which tDCS works are not fully understood, but its primary mechanism of action, which emerges immediately during stimulation involves subthreshold de- or hyperpolarization of neuronal membrane potentials, resulting in excitability-enhancing effects by anodal, and -reducing effects by cathodal stimulation in conventional protocols ^21,22^. In neuropsychiatric disorders that are characterized by functional brain abnormalities (i.e., hyper- or hypoactivity of specific brain region/s and network/s), it is possible to modify altered brain functions with tDCS, and affect target behavior or cognition ^23–26^.

Functional abnormalities of the dorsolateral prefrontal cortex (DLPFC) are well-documented in MD. Specifically, dysbalanced activity between the left and right DLPFC is documented in depression ^27,28^, which has been the rationale for applying the most commonly used tDCS protocol in MD (i.e., anodal left DLPFC, cathodal right DLPFC). In MD, results of tDCS studies have been promising so far ^23,29,30^, and a recent network meta-analysis and an umbrella review of tDCS meta-analyses ^31,32^ further supported its potential for MD treatment. However, an individual participant data (IPD) meta-analysis of nine large RCTs found no difference between active and sham tDCS during the relatively short acute treatment phase but only in the follow-up period ^33^. Nonetheless, there are also studies with negative results or placebo effects of tDCS ^34,35^ that question the efficacy of tDCS for the treatment of MD prompting researchers to focus on optimizing NIBS, including tDCS, interventions. Yet, knowledge about optimal stimulation parameters and the efficacy of interventions is still limited.

The most commonly used tDCS protocol in MD includes applying two relatively large sponge electrodes (e.g., 35 or 25 cm^2^) over the left and right DLPFC, which induces non-focal stimulation of the target regions but also distributes electrical current flow over nearby areas in the brain. It is possible to optimize this protocol using multichannel montages with computational modeling to improve the specificity and enhance the intensity of the induced electrical field in the target region (i.e., left and right DLPFC). In this randomized, double-blind, sham-controlled study, we primarily investigated treatment efficacy of an optimized multi- channel tDCS protocol targeting the left and right DLPFC (using seven small electrodes, maximum of 1.65 mA current per electrode, and a total injected current set to 4.0 mA per polarity), as compared to the conventional bipolar tDCS montage (two 35 cm conventional electrodes, 2 mA max current per electrode and total injected current) on reducing depressive symptoms in 60 patients with MD following 30 sessions of tDCS. Additionally, we investigated treatment response by evaluating symptom changes and monitoring patients over two follow-up assessments (1 month and 3 months). Finally, we investigated how these interventions affect cognitive deficit profiles and affect brain functional connectivity in depression. This study is the first tDCS RCT in MD to examine and compare the impact of a new computationally optimized multi-channel tDCS intervention on symptom reduction in MD with that of conventional (large sponges, bipolar) and sham montages.

## 2. Methods

### 2.1. Participants

This was a randomized, double-blind, parallel-group study. We included patients with unipolar, nonpsychotic MDD per DSM-5 criteria, confirmed by psychiatrists using the Mini-International Neuropsychiatric Interview (MINI). Sixty patients were recruited from several neuropsychiatric clinics across the city of XX from 2021-2023. Patients were randomly assigned to the active multichannel tDCS (n=20), active conventional tDCS (n=20), and sham tDCS (n=20) groups using the block randomization method (block size of 6- see supplementary materials). Sample size was calculated using power analysis based on a small-to-medium effect size (f=0.20 equivalent to *η*p2=0.04, α=0.05, power=0.95, mixed-model ANOVA, *N*=57). We added 3 more subjects to the total sample size to compensate for potential dropouts and increase power. The inclusion criteria were: (1) being 18-60 years of age, (2) failure with at least one antidepressant medication, (3) not currently using antidepressants or having a stable treatment regimen for at least 6 weeks before and throughout the experiment, and (4) unipolar depression diagnosis. Patients had to have a mild to severe depression according to the Hamilton Depression Rating Scale, as well as a low risk of suicide (evaluated based on the MINI). Exclusion criteria were other Axis I disorders, including alcohol or substance use or dependence (anxiety disorders comorbidity was allowed); any Axis II disorders; previous neurological conditions; and specific contraindications for tDCS (e.g. metallic plates in the head). All participants were native speakers and had normal or corrected-to-normal vision. This registered clinical trial was approved by the Ethics Committee of the XXX (XXX.1400.059). Participants provided written informed consent after receiving a complete description of the study. See supplementary Table S1 for basic demographics.

### 2.2. Randomization and blinding

A random allocation sequence table was generated by an online block randomization website (https://www.sealedenvelope.com/simple-randomiser/v1/lists) for three treatment groups and a block size of 6 and 10 blocks to cover 60 patients (patients per group per block: 2). This allocation was carried out with a predetermined block size of 6, ensuring a balanced distribution of participants across the groups. To maintain the integrity of the study and ensure unbiased results, the enroller was blinded to the group allocation, and participant allocation was concealed through a central coordinator (3^rd^ coauthor) who generated and holds the randomization sequence. Patients and intervention codes were securely transferred to the patient’s enroller by the coordinator (via phone call) while the enroller had no access to the block sequence and group assignment. The coordinator had no involvement in the actual study procedure and was merely responsible for randomization coordination. Clinical outcome measures were assessed by a licensed psychiatrist from the hospital, who had no involvement in the study. The investigator responsible for evaluating cognitive functions and functional connectivity was blinded to group assignments. Additionally, a separate experimenter, tasked with measuring outcomes, was unaware of the specific tDCS conditions. To prevent potential bias from unintentional disclosure, patients in each group were kept unaware of one another, ensuring that differences in intervention setups (multichannel cap vs. conventional/sham tDCS) did not compromise blinding.

### 2.3. Clinical outcome measures

The primary outcome measures to examine the effects of the intervention on MD symptoms were the change in depressive symptoms measured by the Hamilton Depression Rating Scale (HDRS-17) score from baseline ^36^ and Montgomery–Åsberg Depression Rating Scale (MADRS) ^37^ score changes versus baseline. The Beck Depression Inventory (BDI-II) ^38^, a self-report questionnaire in contrast to clinician-administered HDRS and MADRS, was also used as the secondary outcome measure. For a detailed description of these measures see supplementary information.

### 2.4. Cognitive assessment

A neuropsychological test dedicated to depression from the CANTAB computerized test battery ^39^ was used to assess cognitive deficits in patients before and after the intervention. The battery includes computerized tasks measuring working memory (Spatial Working Memory- SWM), attention (Rapid Visual Processing- RVP), and executive functions (One Touch Stockings of Cambridge- OTS). A detailed description of these measures is provided in the supplementary information. Briefly, the SWM measures working memory and executive functions. Participants are presented with a series of boxes depicted on a screen. Some boxes contain tokens, and the goal is to find and remember the locations of these tokens ^40^. The RVP is a sensitive measure of sustained visual attention ^41^ and presents participants with a white box in the center of the computer screen, inside which digits from 2 to 9 appear in a pseudo-random order at the rate of 100 digits per minute. Participants are asked to detect target sequences of numbers and respond using a press pad. The OTS, a modified version of the Tower of London task, requires participants to rearrange colored balls in vertical columns to match a desired final arrangement in a specified minimum number of moves ^40^.

### 2.5. Brain functional connectivity

#### 2.5.1. EEG Preprocessing and Artifact Removal

EEG signals were recorded with a 21-channel EEG device (Mitsar, Ltd, Russia) with a sampling rate of 250 Hz, and the included electrodes were Fz, Cz, Pz, C3, T3, C4, T4, Fp1, Fp2, F3, F4, F7, F8, P3, P4, T5, T6, O1, O2, A1, A2. The recordings were obtained in the eyes-open condition for three minutes and then in eyes-closed condition for three minutes (six minutes in total). To preprocess and remove artifacts from the EEG data, we used Makoto’s preprocessing pipeline in the EEGLAB toolbox 2022.1 ^42^ with MATLAB 2022b (The MathWorks, Natick, MA). The data were first resampled to 512 Hz, high-pass filtered at 1 Hz and re-referenced to an average reference. We used the CleanlineNoise plugin in EEGLAB for line noise removal. We then applied ASR (Artifact Subspace Reconstruction), an automated algorithm that eliminates flatline and noisy channels, low-frequency drifts, and short-time bursts. Any removed channels were interpolated using the spherical interpolation method. Afterward, we visually inspected all the raw data to detect artifact-related parts. To remove non-brain artifacts, we applied Adaptive Mixture ICA (AMICA) to the EEG data to decompose independent components (ICs). We then used the EEGLAB plugin ICLabel to identify brain ICs (with a ’brain’ label probability of more than 0.8) from all types of ICs, including Brain, Muscle, Eye, Heart, and others. Finally, we extracted a 60-second segment from the middle part of each preprocessed EEG and exported it to the BRAINSTORM software (version November 2024) for further analysis ^43^.

#### 2.5.2. EEG analysis

Functional connectivity was assessed using multiple metrics to capture distinct aspects of neural synchronization. The time series from each EEG channel was segmented into 60-second epochs and bandpass filtered into seven frequency bands (delta: 1–4 Hz, theta: 4–8 Hz, alpha 1: 8–10 Hz, alpha 2: 11–13 Hz, beta 1: 13–21 Hz, beta 2: 19–30 Hz, gamma: 30–40 Hz) with finite impulse response filters. The Hilbert transform was then applied to each filtered signal to extract its instantaneous phase. For each pair of EEG channels, we quantified connectivity using three complementary methods: First, we calculated the Phase-Locking Value (PLV) ^44^ which is sensitive to the synchronization of oscillatory neural activity and reflects dynamic coordination across brain networks. In addition, we computed the Magnitude-Squared Coherence ^45^, capturing both phase and amplitude relationships by correlating the power spectra of two signals. Lastly, we analyzed global efficiency, a measure of functional connectivity associated with information flow and cognitive performance, using the Weighted Phase Lag Index (wPLI) ^46^. These connectivity measures were chosen as previous studies have shown that depression is generally associated with reduced and/or altered synchronization patterns ^10,11^ and lower coherence ^11,12^ in the default mode network (DMN) and central executive network (CEN). Further details about these measures, EEG data preprocessing, and analysis are provided in the supplementary information.

### 2.6. tDCS

Two electrical stimulators for feasibility due to the high number of daily sessions per subject. To test two tDCS devices for a constant 2 mA output, we connected a 5 kΩ load resistor across each device’s electrodes to simulate human head resistance. We used a digital multimeter set to DC current mode, placed it in series with the resistor, and powered on each device, measuring the current to confirm it stabilized at 1.9–2.1 mA. Also, we used an oscilloscope in DC coupling mode to check for noise or ripples across the resistor, ensuring a clean DC output. We repeated the process for both devices, comparing their performance under identical conditions. The electrode texture was the same in all stimulation conditions. In conventional tDCS, direct currents were generated by an electrical stimulator and applied through a pair of saline-soaked sponge rubber electrodes (7×5 cm) for 30 minutes (with 30 s ramping up and down) and with an intensity of 2 mA. In sham tDCS, the electrical current was ramped up and down for 30 seconds and then turned off in the beginning of study and repeated at the end of stimulation to mimick active tDCS sensations in patients ^47^. In the group-optimized multichannel protocols, stimulation was delivered with the StarStim 8 stimulator (Neuroelectrics, Spain) using 7 small electrodes whose positions and currents were determined with the Stimweaver algorithm^48^. A side-effect survey was done after each tDCS session ^49^. To avoid participant bias potentially due to habituation to tDCS-induced sensations over multiple sessions, blinding efficacy was not assessed among patients at the study endpoint. However, following the second follow-up at three months, all patients were asked to select one of three options (0, 1, 2), representing no stimulation, moderate stimulation, and strong stimulation, respectively. Patients in the sham group were subsequently offered an active tDCS intervention (conventional) after being informed about the sham condition. This phase was not included in the original study design.

### 2.7. Procedure

First, participants completed a brief questionnaire to evaluate their suitability for brain stimulation. All participants received 30 sessions of stimulation over 5 consecutive weeks (1 session daily from Saturday to Friday, 5 days per week). To avoid confounding effects of the intervention at a circadian non-preferred time of day, which can significantly affect neuroplasticity induction ^50^, all stimulation sessions took place between 11:00-14:00, and participants were not under sleep pressure ^51^. Clinical outcome measures were evaluated before the first intervention (pre-intervention-T0), after the 10^th^ session (T1), after the 20^th^ session (T2), after the last session (post-intervention-T3), 1 month (T4), and 3 months (T5) following the last stimulation session. Cognitive function assessment and EEG recordings took place only before and after the intervention. Patients were instructed about cognitive tasks before the beginning of the experiment. None of the patients underwent psychotherapy at any point during the study.

Participants were unaware of the study hypotheses and stimulation conditions. To ensure treatment adherence and attendance of assessment sessions, all participants were offered free transportation for assessment sessions and could choose to complete the clinical evaluation remotely in case COVID-19 restrictions compromised the patients’ willingness.

### 2.8. Statistical analysis

Clinical and behavioral data analyses were conducted with the statistical package SPSS, version 26.0 (IBM, SPSS, Inc., Chicago, IL), and GraphPad Prism 8.2.1 (GraphPad Software, San Diego, California). The normality and homogeneity of data distribution, and variance were confirmed by Shapiro-Wilk and Levene tests, respectively. Between-group differences in demographic variables were explored by Chi-square tests or Fisher’s exact test for categorical variables and F-tests for continuous variables. Mixed model ANOVAs were conducted for the dependent variables with “group” (multichannel tDCS, conventional tDCS, sham tDCS) as the between-subject and time (five timepoints for clinical measures: pre-intervention, T1, T2, post-intervention, 1-month follow-up, 3-month follow-up; two timepoints for cognitive/EEG measures) as the within-subject factors. Mauchly’s test was used to evaluate the sphericity of the data before performing the respective ANOVAs. In case of violation, degrees of freedom were corrected using Greenhouse-Geisser estimates of sphericity. *Post hoc* analyses were calculated using Tukey’s multiple comparisons tests to examine individual mean difference comparisons across groups (active multichannel, active conventional, sham) and time points (pre-intervention, session 10, session 20, session 30, 1-month follow-up, and 3-month follow-up). Pearson’s correlational analyses explored associations between reported side effects and study outcome measures. The critical level of significance was 0.05 for all statistical analyses. For functional connectivity, first within-group differences in post- versus pre-intervention connectivity were evaluated with non-parametric permutation paired t-tests in Brainstorm ^43^, set at 8,000 iterations (*p*≤0.05), and applying FDR correction, targeting each connection in the connectivity matrices.

Next, between-group differences in pre-intervention connectivity values were assessed using non-parametric permutation-independent t-tests with the same parameters. Finally, between-group differences in post-intervention connectivity were examined using the same approach. All between-group analyses were conducted to identify significant differences in functional connectivity across experimental conditions.

## 3. Results

### 3.1. Data overview

Participants tolerated the stimulation well, and no serious adverse effects were reported during or after the intervention (Table S2). The optimized multichannel tDCS group reported more itching, tingling, and burning sensations during stimulation, along with increased skin redness, headaches, concentration difficulties, and sleep issues afterward, compared to the sham group. The conventional tDCS and sham groups experienced higher levels of fatigue after the intervention compared to the multi-channel tDCS group. Despite the higher reported side effects in the active tDCS groups, the Pearson correlational analyses provide evidence that these side effects generally did not influence the clinical outcome measures (HDRS and MADRS) (supplementary content) suggesting that the higher side effects in the multichannel and conventional tDCS groups did not substantially affect the clinical efficacy of the treatments. No significant differences were observed in demographic data (Table S1) or baseline clinical/cognitive outcomes among the three groups (Table S3).

### 3.2. Primary clinical measures: Intervention efficacy and treatment response

The results of the 3 (multichannel tDCS, conventional tDCS, sham tDCS) × 6 (pre, T1, T2, T3, 1-month follow-up, 3-month follow-up) mixed ANOVA revealed significant main effects of time (*F_2.98_*=37.90, *p*<0.001, *η*p^2^=0.39), group (*F_2_*=5.24, *p*=0.008, *η*p^2^=0.15) and importantly group×time interaction (*F_5.96_*=9.34, *p*<0.001, *η*p^2^=0.24) for the HDRS scores.

Tukey’s multiple test comparisons show a significant decrease in HDRS scores at all time points (i.e., session 10, session 20, session 30, 1-month follow-up, and 3-month follow-up) in the multichannel tDCS group as compared to the pre-intervention score. In the conventional tDCS, HDRS scores were significantly reduced only after session 30 when compared to pre-intervention scores. When compared to the sham group, only the optimized multichannel tDCS group showed a significant reduction of HDRS scores at session 20, session 30, 1-month follow-up, and 3-month follow-up assessment and was significantly larger than the conventional tDCS group at session 30, 1-month follow-up and 3-month follow-up assessment. Baseline (pre-intervention) between-group comparisons (active groups vs sham) showed no significant differences (Fig 3A).

**Fig. 1:**
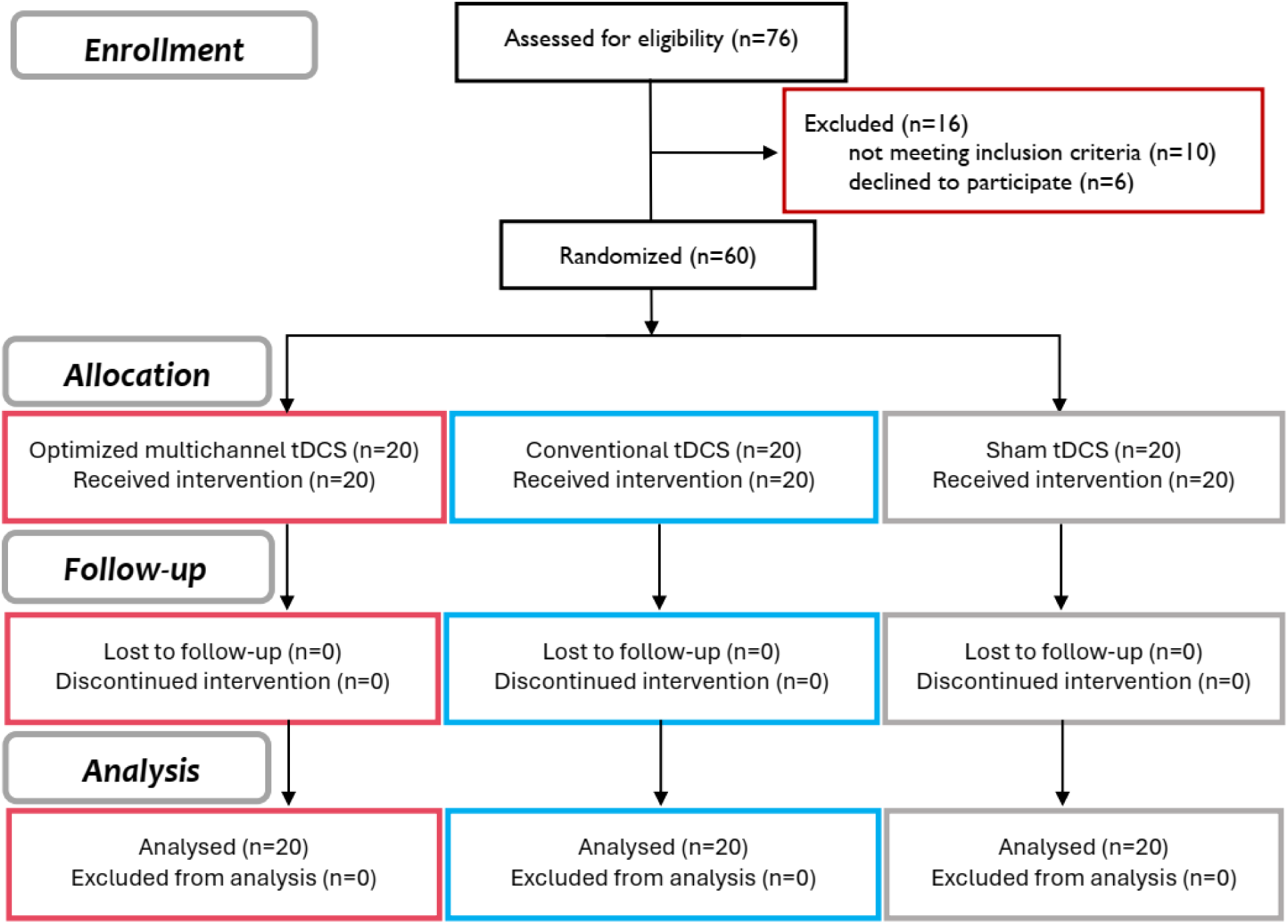
CONSORT diagram of study inclusion.

**Fig. 2:**
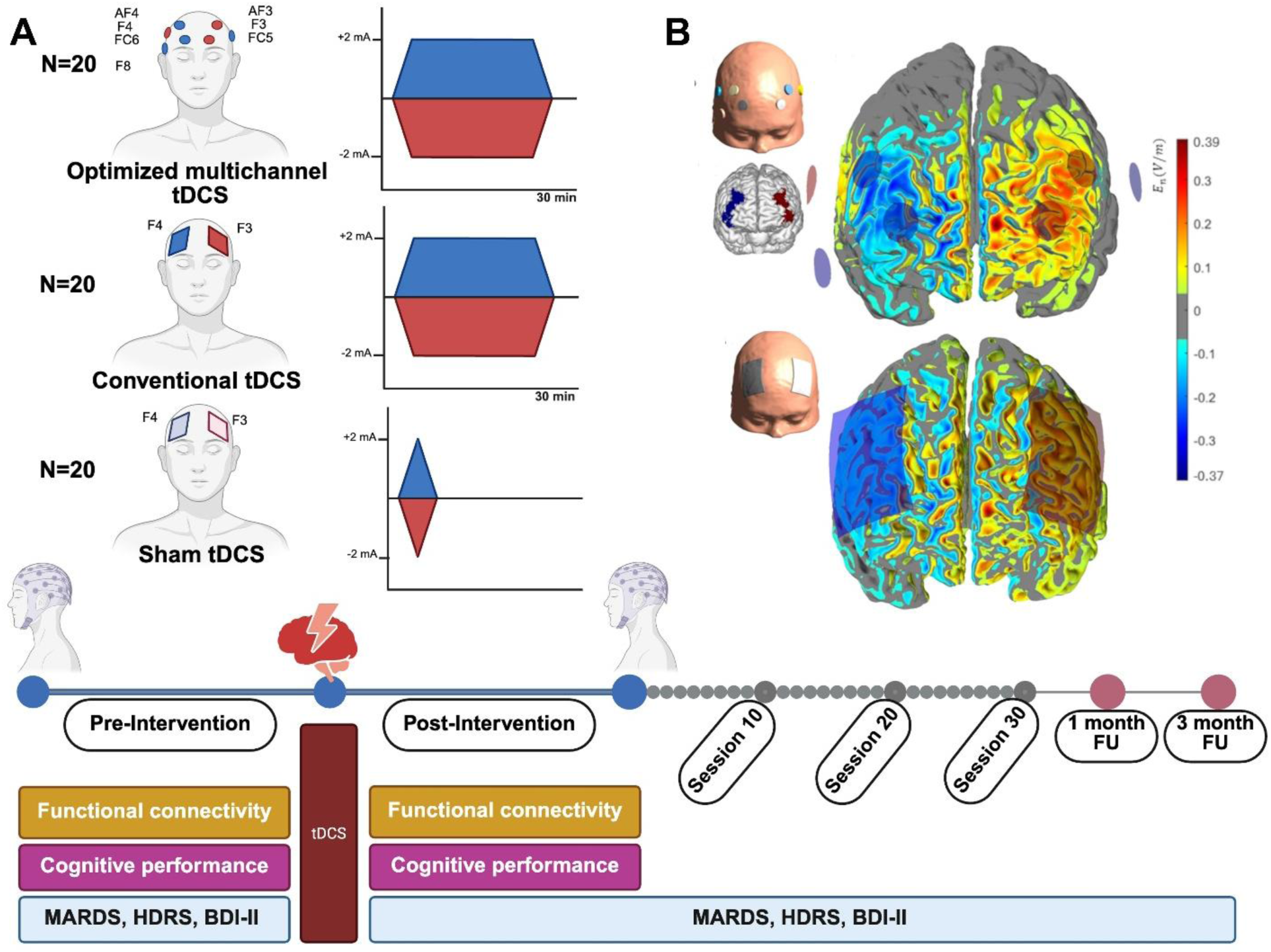
**A**, Study procedure. The experiment was conducted in a randomized, double-blind, sham- controlled parallel-group design. Patients were assigned to one of three tDCS groups: 2-mA conventional tDCS [total injected current 2 mA] (n = 20), 2-mA optimized multichannel tDCS [total injected current 4 mA] (n = 20), and sham tDCS (n = 20). All participants underwent 30 daily stimulation sessions. Depressive symptoms were evaluated before the intervention and up to three months afterward. Cognitive performance and cerebral functional connectivity were assessed only before and after the intervention. **B**, Distribution of the normal component of the E-field (En, in V/m) in a surface inside the grey matter tissue, in between the grey matter-CSF and white matter-grey matter interfaces. Bottom panel shows the En-distribution for a conventional protocol using a bipolar electrode montage with rectangular 7x5 cm^2^ sponge electrodes (anode/cathode over F3/F4) at +2/-2-mA. Top panel illustrates the En-distribution for the group-optimized multichannel protocol using gelled electrodes (1 cm radius PiStim electrodes, anodes shown in red, cathodes in blue) targeting Brodmann area 46, presumed to result in excitability enhancement in the left hemisphere and excitability reduction in the right. Positive En values indicate that the En-field is directed into the cortical surface, thus leading to an increase in cortical excitability, according to the lambda-E model. Negative En values are directed in the opposite direction, thus leading to a decrease in excitability.

**Fig. 3:**
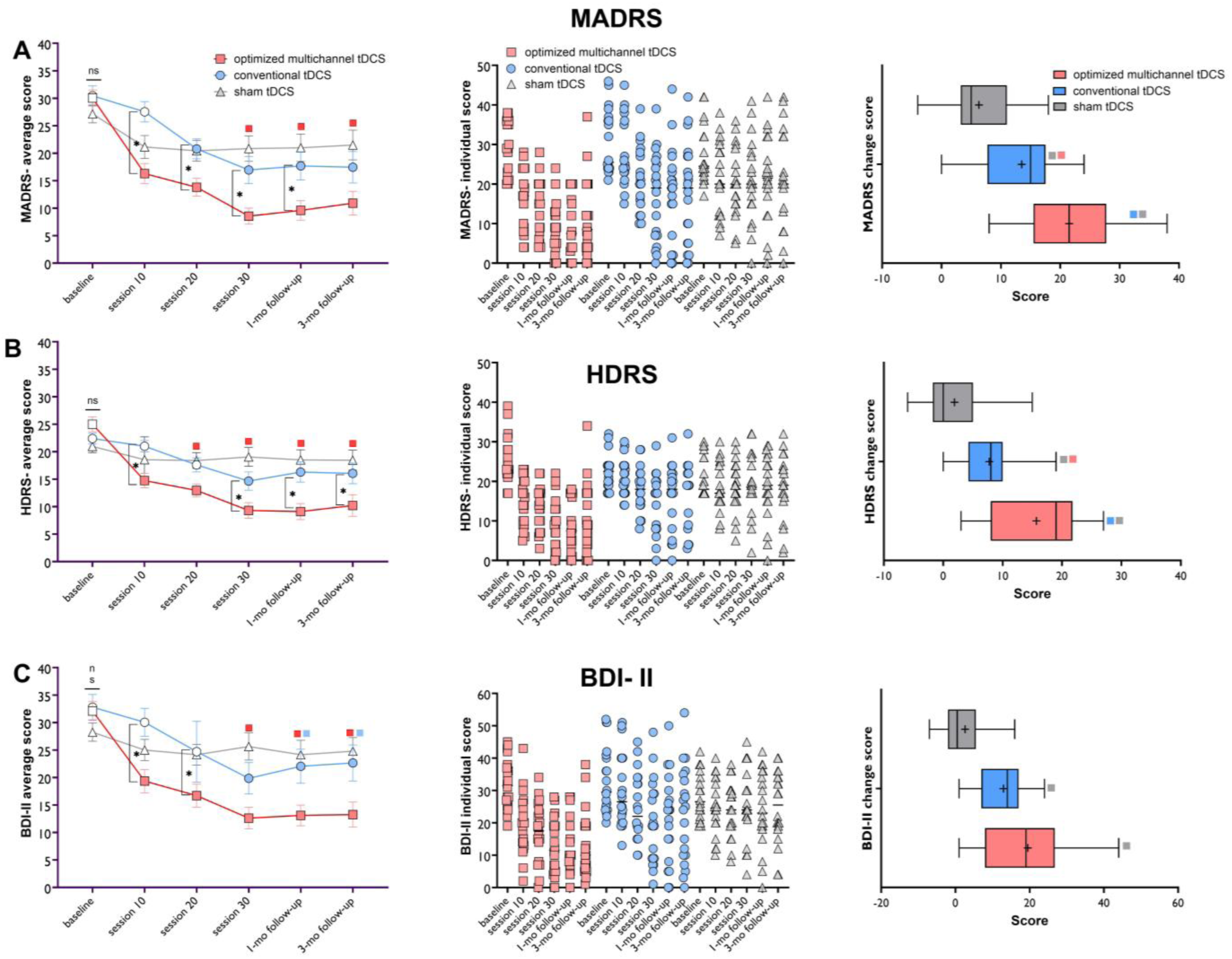
Depressive symptoms measured by the Hamilton Depression Rating Scale (A), Montgomery– Åsberg Depression Rating Scale (B), and Beck Depression Inventory (C) before, after, and up to 3 months following the intervention. In the left panel, filled symbols indicate significant differences at each time point compared to pre-intervention scores. The red and blue floating symbols represent significant differences between the active stimulation groups (conventional and multichannel tDCS) and sham tDCS at each time point. Asterisks show the difference between the multichannel vs sham tDCS groups at respective time points. The middle panel displays a scatter plot of clinical scores for each group across time points. The right panel shows the mean score change from baseline to the study endpoint (week 6) for the respective measure. The horizontal bar shows the median, the + shows the mean, the upper and lower boundaries show the 25th and 75th percentiles, respectively, and the whiskers show 1–99 percentile. All pairwise comparisons are conducted with Tukey’s multiple comparisons test, and error bars indicate the standard error of the mean (s.e.m.). *Note*: HDRS = Hamilton Depression Rating Scale; MADRS = Montgomery–Åsberg Depression Rating Scale; BDI-II = Beck Depression Inventory-II; tDCS = transcranial direct current stimulation; mo = month.

Similarly for MADRS scores, a significant main effect of time (*F_3.26_*=57.34, *p*<0.001, *η*p^2^=0.50), group (*F_2_*=5012, *p*=0.008, *η*p^2^=0.15) and also significant group×time interaction (*F_6.53_*=7.51, *p*<0.001, *η*p^2^=0.21) were found. The MARDS scores significantly decreased from session 20 until the last assessment in both multichannel and conventional tDCS groups when compared to the pre-intervention score and no significant changes in the sham group. Compared to the sham group, MARDS scores were significantly reduced only in the multichannel tDCS group at sessions 30, 1-month, and 3-month follow-up assessments and at sessions 10, 20, 30, and 1-month follow-up as compared to the conventional tDCS (Fig 3B).

### 3.3. Secondary clinical measure

The results showed a significant main effect of time (*F_3.14_*=32.15, *p*<0.001, *η*p^2^=0.36) and group (*F_2_*=4.74, *p*=0.012, *η*p^2^=0.14) and group×time interaction (*F_6.29_*=6.10, *p*<0.001, *η*p^2^=0.17) for the BDI-II scores. Tukey’s multiple test comparisons show that the multichannel tDCS significantly reduced BDI-II scores at all time points as compared to the pre-intervention score while the conventional tDCS group significantly reduced BDI-II scores at session 30 and both follow-up measurements (Fig 4C). No significant score reduction was found in the sham group after the intervention. When compared to the sham group, BDI-II scores were significantly decreased in the multichannel tDCS group at sessions 30, 1-month, and 3-month follow-up assessments and in the conventional tDCS group at both follow-up assessments. The BDI-II scores in the multichannel group were also significantly lower at sessions 10 and 20 as compared to the conventional tDCS (Fig 3C).

**Fig. 4:**
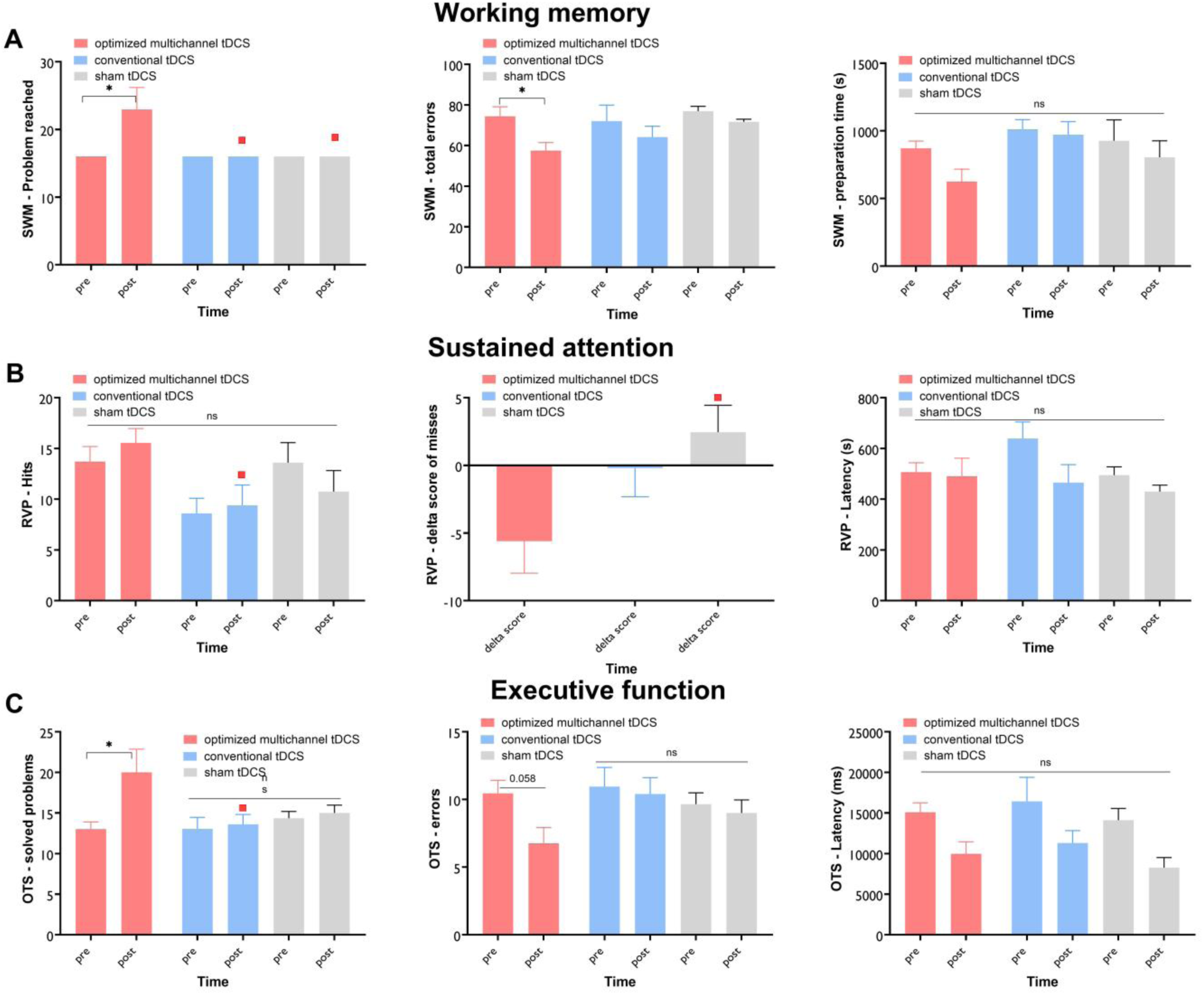
Cognitive performance was evaluated before and after 30 sessions of optimized multichannel, conventional, and sham tDCS using the CANTAB neuropsychological test battery for depression, which includes measures of working memory (SWM) (A), sustained attention (RVP) (B), and executive functioning (OTS) (C). Asterisks indicate significant differences for pre- and post-intervention comparisons while floating symbols, depending on the color, denote significant group differences (in this case multichannel tDCS vs sham or conventional tDCS) at each time point. Tukey’s multiple-test method was used for all pairwise comparisons, and error bars indicate the standard error of the mean (s.e.m.). Note: SWM = Spatial Working Memory; RVP = Rapid Visual Processing; OTS = One Touch Stocking of Cambridge; tDCS = transcranial direct current stimulation.

### 3.4. Cognitive performance

Working memory, sustained attention, and executive functions were assessed before and after the intervention with the SWM, RVP, and OTS tasks respectively. For working memory, the 3×2 mixed model ANOVA results revealed significant main effects of time (*F*=4.85, *p*=0.032, *ηp^2^*=0.080), group (*F*=4.77, *p*=0.012, *ηp^2^*=0.146), and group×time interaction (*F*=4.73, *p*=0.012, *ηp^2^*=0.146) on the number of solved problems. Tukey’s multiple comparisons show that only the optimized multichannel tDCS significantly increased the number of solved problems compared to sham (*p*=0.025) and conventional tDCS (*p*=0.025) and also compared to the pre-intervention measure (*p*<0.001). A significant main effect of time was observed on performance error (*F*=14.23, *p*<0.001, *ηp^2^*=0.200), but effects of group or group×time interactions. Post-hoc comparisons of post-intervention vs pre-intervention error rates show a significant error reduction after optimized multichannel tDCS (*p*=0.039, -22.7%) but not in the conventional (-11.51%) and sham tDCS conditions (-3.69%). For the preparation time to response, only a significant main effect of time was found (*F*=5.49, *p*=0.023, *ηp^2^*=0.089).

Tukey’s multiple test comparisons of post-intervention vs pre-intervention scores, however, showed no significant decrease in preparation time after the optimized multichannel (-28.2%), conventional (-3.98%), and sham tDCS (-14.4%) (Fig 4A).

For sustained attention, the 3×2 mixed ANOVA revealed significant main effects of group on the number of hits (*F*=4.15, *p*=0.021, *ηp^2^*=0.136) and misses (*F*=5.21, *p*=0.010, *ηp^2^*=0.150), along with a significant group×time interaction effect on misses (*F*=3.58, *p*=0.034, *ηp^2^*=0.112) (Table 3). Following post-hoc comparisons, no significant changes were observed in hits or misses post-intervention compared to pre-intervention scores in either group. The number of hits after the intervention was, however, significantly higher in the multichannel tDCS as compared to the conventional, but not sham tDCS. Moreover, delta scores (post-intervention score minus pre-intervention score) were analyzed using a univariate ANOVA. The results showed a significant main effect of group (*F*=3.58, *p*=0.034, *ηp^2^*=0.112), and Tukey’s post hoc comparison indicated that missed trials were significantly lower in the optimized multichannel tDCS group than in the sham tDCS group (*p*=0.029). Regarding performance speed (mean latency), significant main effects of time (*F*=5.97, *p*=0.019, *ηp^2^*=0.130) and group (*F*=4.09, *p*=0.024, *ηp^2^*=0.170) were observed, but no significant group×time interaction was found. Post hoc tests revealed no significant differences in performance speed within or between groups (Fig 4B).

Finally for executive functioning, the results of 3×2 mixed ANOVA revealed a significant main effect of time (*F*=7.30, *p*=0.009, *ηp^2^*=0.114) and a group×time interaction (*F*=4.51, *p*=0.016, *ηp^2^*=0.135) on the number of solved problems. For the number of errors before the correct response (*F*=7.66, *p*=0.008, *ηp^2^*=0.119) and the mean time taken to reach a correct response (*F*=19.82, *p*<0.001, *ηp^2^*=0.258), there was also a significant main effect of time. Tukey’s multiple comparisons indicated that only the optimized multichannel tDCS group showed a significantly higher number of solved problems post-intervention compared to pre-intervention (*p*=0.004). No significant differences were found between the active and the sham stimulation protocols in terms of committed errors and performance time, although the error reduction was numerically greater after optimized multichannel tDCS (-35.3%) than the conventional (-5.02%) and sham tDCS interventions (-6.73%) (Fig 4C).

### 3.5. Functional connectivity

Brain functional connectivity changes before and after the intervention were measured with key connectivity measures related to cognitive impairment and emotional regulation such as PLV, coherence, and global efficiency. Significant changes were only observed in the post-intervention connectivity of the optimized multi-channel tDCS compared to the sham tDCS (Figure 6). Specifically, theta, alpha, and beta PLV values significantly increased in the left frontal-right temporal and left and right occipital regions following optimized multi-channel tDCS compared to sham tDCS. For coherence, significant increases were observed in theta, alpha, and beta coherence values after the optimized multi-channel tDCS across the left frontal and right temporal regions, as well as for prefrontal-central coherence. Additionally, global efficiency measured by wPLI showed a similar increase in connectivity within the prefrontal- central region in the delta band and the Fronto-Parietal Network in the gamma band (see Figure 5 in supplementary for details).

## 4. Discussion

In this randomized, sham-controlled clinical trial, we evaluated the efficacy of optimized multichannel and conventional tDCS protocols compared to sham tDCS over the prefrontal cortex on clinical symptoms, cognitive deficits, and cerebral functional connectivity in patients with MD. Both optimized multichannel and conventional tDCS significantly alleviated depressive symptoms immediately after the intervention (session 30), and these improvements were sustained for up to three months post-intervention, outperforming the sham group.

Therapeutic effects emerged as early as session 10 in the multichannel tDCS group, with the optimized multichannel protocol demonstrating a significantly greater effect size than conventional tDCS. Furthermore, the optimized multichannel tDCS group showed superior enhancement in cognitive performance, particularly in working memory and executive functions, compared to both conventional and sham tDCS. This protocol also led to increased cerebral functional connectivity, as evidenced by enhanced PLV, coherence, and global efficiency metrics, following the intervention. In contrast, no significant changes were observed in the sham group across outcome measures. This study is one of the largest tDCS trials conducted for depression treatment and is the first to utilize an optimized multi-channel tDCS protocol with a total stimulation intensity of 4 mA. In what follows we primarily discuss the clinical efficacy of the intervention, the study’s primary objective, and the implications of cognitive and cerebral connectivity improvements for these findings.

### 4.1. Clinical efficacy

The study primarily aimed to assess the clinical efficacy of optimized multichannel tDCS and conventional tDCS versus sham tDCS. We found that optimized multichannel tDCS was clinically superior to both conventional and sham tDCS, although conventional tDCS also effectively reduced depressive symptoms compared to sham. Although the therapeutic effects of tDCS in depression, compared to other neuropsychiatric disorders, are well-supported in previous ^23,29^ and recent well-conducted studies ^52,53^, its clinical efficacy is not yet established and studies with negative and/or placebo effects ^35,54^ have questioned tDCS efficacy. Comparing the clinical efficacy of this study to recent studies is, nevertheless, informative. One of the largest tDCS studies for depression treatment investigated the efficacy of conventional home-used tDCS^52^. In this study, which employed a conventional tDCS protocol similar to ours, the active tDCS group showed a significant improvement from baseline to week 10, with a mean reduction of 11.31 points on the MADRS compared to 7.74 points for sham treatment. In another study, remotely supervised multichannel tDCS was conducted at home in MD patients (N=35) ^55^. The Ruffini et al. study reported a median MADRS score reduction of 19.8 points (64.5%; 48.6, 72.4) four weeks post-treatment, with Hedge’s g = 3.10. In our study, we observed a response rate of 72.7% (n=24) with at least a 50% improvement in MADRS scores from baseline to the last visit four weeks post-treatment (Hedge’s g = 2.80). Other clinical measures in our study indicated similar enhancements, with a reduction of about 12 points in the conventional group, 19 points in the multichannel active group, and 8 points in the sham group. Our results align well with studies using conventional and multichannel tDCS at home in terms of symptom reduction after conventional and multichannel tDCS. Additionally, they indicate that the optimized multichannel tDCS demonstrates greater clinical efficacy, achieving approximately a 19-point reduction in the MADRS compared to 13.5 points in conventional and 6.3 points in sham tDCS. The active protocols’ overall efficacy aligns with recent meta-analyses indicating that a 2-mA stimulation current during a 30-minute tDCS session can enhance intervention effectiveness ^56^. However, our study revealed significantly greater efficacy when this intervention is applied using an optimized multichannel configuration, which will be discussed further.

Furthermore, this finding can be partially explained from a neurophysiological perspective. The hallmark finding of neuroimaging studies refers to a lateral hypoconnectivity in the prefrontal cortex (especially in the left DLPFC) ^13,27^, which is the rationale for applying the most commonly used stimulation protocol in depression, excitatory left inhibitory right DLPFC stimulation ^30^. We applied anodal and cathodal stimulation over the left and right DLPFCs to un and downregulate the activity of these regions respectively. With causal modulation of cortical excitability with tDCS ^14^, we expected to restore functional abnormalities in the MD-relevant brain circuitry, and this intervention was associated with behavioral and clinical improvement in this study. Moreover, clinical efficacy of the multichannel optimized stimulation was observed from the 10^th^ session up to 3-month follow-up, which was not the case in the conventional tDCS and sham groups. The protocol used in the optimized multichannel group was designed by computational modeling ^48^ to deliver an En-field distribution that closely fits the target map, aiming to excite the left DLPFC and inhibit the right DLPFC. This optimization resulted in a significantly better fit of the multichannel En-field distribution to the target map, as reflected in the much higher NERNI (a normalized version of ERNI) value of 0.17 for the multichannel protocol compared to 0.018 for the bipolar protocol. This improved fit aligns with the larger effect size of clinical measures observed in the multichannel tDCS group and cognitive enhancement. Additionally, the optimized multichannel protocol allowed for higher total injected currents (4.0 mA, double that of the 2 mA bipolar protocol) which might partially explain larger clinical efficacy and cognitive outperformance in the optimized multichannel tDCS. In the template head model used for optimization, this resulted in an average En-field on target that was 1.3-1.4 times larger (for the left and right target areas, respectively) in the multichannel montage compared to the bipolar montage. Both the fit to the target map and the E-field dose have been shown to correlate with quantitative measures of stimulation effects in other applications ^57,58^.

Furthermore, other trials have demonstrated the effectiveness of using a modeling approach to optimize stimulation dose parameters in depression, even when employing variations of the target map used in this study ^55^.

### 4.2. Cognitive performance and brain functional connectivity

The optimized multichannel tDCS significantly enhanced cognitive performance, specifically working memory and executive functions, in patients following the intervention, unlike conventional and sham tDCS. Notably, these cognitive deficits are key symptoms of MD, and their improvement, alongside clinical symptoms, indicates that the optimized tDCS intervention positively impacted both mood and cognitive deficits in MD, which are interconnected ^59,60^. These cognitive deficits were repeatedly shown in MD ^61^ but results about cognitive effects of tDCS in depression and other disorders have been mixed or limited to memory functioning ^26,62^ and our study supports the cognitive potency of tDCS in MD ^26^.

The changes in brain functional connectivity measures after the optimized multichannel vs sham tDCS protocol are in line with reduced biomarkers of depression symptoms. Previous EEG studies in depression have shown reduced synchronization (measured by PLV) across regions, specifically in the DMN, and prefrontal-limbic circuits ^10,11^ which are linked to emotional dysregulation, and cognitive impairment in depression. Similarly, lower coherence in frontotemporal regions and across theta, alpha, and beta bands has been observed in patients with depression. Our results indicate significant increases in PLV and coherence in these areas and frequency bands (see Figure 6), suggesting that targeted brain regions exhibit enhanced functional connectivity and synchronous cerebral oscillatory activity (e.g., theta and alpha bands) after the optimized multichannel tDCS intervention. These changes are associated with improved core cognitive functions (i.e., working memory and executive control) and regulated emotions (measured by clinical measures), which are usually seen in depression treatment response ^59,60,63^.

### 4.3. Conclusion

Findings of this study show superior clinical efficacy and cognitive effects of the optimized multi-channel tDCS vs conventional and sham tDCS for treating depression. This study highlights the importance of optimizing stimulation parameters to effectively target specific cortical regions in the brain. Additionally, it suggests that optimized multichannel tDCS could be more effective in treating other neuropsychiatric disorders, and suggests the value of further research using computational modeling for precision stimulation ^64^. The findings also underscore the significance of session frequency in assessing clinical efficacy, particularly in conventional tDCS protocols. Notably, the conventional tDCS significantly reduced depressive symptoms in two clinical outcome measures (HRDS and BDI-II) only after the 30th session. At the same time, the optimized multichannel tDCS showed reductions in depressive symptoms from the 10^th^ session across all measures. This study had several limitations including the inability to assess cognitive functions and resting EEG during follow-up due to COVID-19 and technical issues. Longer-term data could have provided deeper insights into tDCS effects on brain physiology and neuropsychological performance. We also did not evaluate blinding efficacy by asking patients about their intervention type. While stimulation intensity may lead to different skin sensations, reported tDCS side effects were similar across groups. Nonetheless, our findings support the effectiveness of the optimized multichannel tDCS for major depression and potentially other neuropsychiatric disorders, with improvements in treatment-related variables noted, especially in the optimized group, enhancing our understanding of treatment efficacy.

## Data Availability

All data generated in this study will be made available upon reasonable request to the authors following the publication of a peer-reviewed version of the manuscript.

---

**Table S1.** Demographic data.

|  |  | Optimized<br>multichannel tDCS | conventional<br>tDCS | sham<br>tDCS | *p* * |
| --- | --- | --- | --- | --- | --- |
| Sample size (*n*) |  | 20 | 20 | 20 |  |
| Age – Mean (SD) |  | 32.15 (12.18) | 35.75 (11.38) | 29.60 (8.06) | 0.197 |
| Sex – Male (female) |  | 11 (9) | 11 (9) | 5 (15) | 0.089 |
| Job | student | 7 | 14 | 10 | 0.241 |
|  | self-employed | 5 | 5 | 3 |  |
|  | housewife/husband | 3 | 7 | 1 |  |
|  | employee | 4 | 3 | 6 |  |
|  | unemployed | 1 | 1 | 0 |  |
| Education | Below diploma | 1 | 5 | 0 | 0.047 |
|  | Diploma | 10 | 7 | 4 |  |
|  | Bachelors degree | 6 | 4 | 11 |  |
|  | Masters degree | 1 | 2 | 5 |  |
|  | PhD level | 2 | 2 | 0 |  |
| Marital status | single | 9 | 11 | 12 | 0.457 |
|  | married | 10 | 9 | 7 |  |
|  | divorced | 1 | 0 | 1 |  |
| On medication (n) |  | 17 | 18 | 15 | 0.368 |
| Type of medication | SSRI | 14 | 16 | 13 |  |
|  | Tricyclic | 3 | 2 | 2 |  |
|  | Other | 0 | 0 | 0 |  |

*Note:* tDCS = transcranial Direct Current Stimulation; M = Mean; SD = Standard Deviation. \* = between-group differences in demographic variables were explored by Chi-square tests or Fisher's exact test for categorical variables and *F* tests for continuous variables.

